# Response of the Myocardium to Hypertrophic Conditions in the Adult Population: The Prospective Observational REMODEL Cohort Study Protocol

**DOI:** 10.1101/2025.05.04.25326959

**Authors:** Vivian Lee, Troy Puar, Michelle SL Kui, Jennifer A Bryant, Yiying Han, Aisyah Binte Latib, Warren Tay, Siew Ching Kong, Desiree-Faye Toh, Redha Boubertakh, Chi-Hang Lee, Thu-Thao Le, Calvin WL Chin

## Abstract

**Background:** In addition to hypertension, the constellation of metabolic abnormalities (diabetes mellitus, dyslipidemia and/or obesity) independently increases the incidence and severity of cardiovascular diseases, and this is compounded by the modern lifestyle and aging society. The prevalence of metabolic syndrome is high and non-hypertensive heart failure is common in Asians. Adverse cardiac remodeling is an important substrate for cardiac dysfunction in the onset and progression of heart failure and its amelioration improves outcomes and prognosis. Better understanding of metabolic-driven cardiac remodeling is warranted due to the rising prevalence and complexity of metabolic syndrome and strong interests in targeted therapy.

**Methods and Analysis:** Response of the Myocardium to Hypertrophic Conditions in the Adult Population (REMODEL) is a prospective observational cohort study with an aim to establish the significance of cardiac remodeling by cardiovascular magnetic resonance (CMR). Expanding from the initial hypertension population, the study examines adults with cardiometabolic conditions including hypertension, diabetes, dyslipidemia, obesity and fatty liver disease. Cardiac remodeling features pertaining to hypertrophy, fibrosis and functional changes are assessed on CMR. Body adiposity is mapped by magnetic resonance imaging across the heart, liver and abdomen. Outcome data is adjudicated and follow-up assessment will be available in a subset of participants. Blood biomarkers will be investigated in relation to imaging findings. Cross-sectional analysis will establish the implication of cardiometabolic disease towards cardiac remodeling, while follow-up and outcome analysis will infer on disease progression and prognosis.

**Ethics and Dissemination:** The study was approved by the SingHealth Centralised Institutional Review Board (2015/2603). Study outcomes will be published in peer-reviewed journals and scientific conferences.

**Trial Registration Number:** NCT02670031

**STRENGTHS AND LIMITATIONS OF THIS STUDY:**

- This prospective cohort study will allow targeted interrogation of the interplay between metabolic disorders and cardiac remodeling in a population-based multiethnic Asian cohort.
- Longitudinal and biomarker analyses will elucidate cardiac remodeling progression and identify early and novel cardiometabolic risk markers.
- Self-reporting data such as lifestyle habits and family history may be subject to recall bias.

## INTRODUCTION

Cardiovascular diseases constitute the greatest cause of global morbidity and mortality, of which, 17% is attributed to metabolic risk factors. The prevalence of metabolic disease is approximately double in Singapore at 34%[1].

The heart as an energy-demanding organ relies on highly regulated metabolism to meet its physiological needs. As such, systemic and cardiac metabolism derangement underlies many cardiovascular remodeling and pathologies, giving rise to cardiometabolic disorders. Often present together, major metabolic risk factors in patients with cardiovascular diseases include hypertension, hyperglycemia and obesity. In hypertensive heart disease, pressure overload on the heart as a result of chronic elevated blood pressure induces left ventricular hypertrophy (LVH) to compensate the increase in ventricular wall stress[2]. Meanwhile, evidence of adverse cardiac remodeling accompanied by myocardial fibrosis is more apparent in patients with diabetes mellitus than those without[3,4]. Similarly, dysregulated adiposity at ectopic sites such as the visceral compartments at the abdomen, heart and liver is associated with adverse cardiac remodeling[5]. The tendency of co-existence and interplay among these metabolic risk factors further complicate the pathogenesis, and thus treatment and prevention of cardiometabolic diseases.

Despite extensive association studies, the pathophysiology and prognosis of adverse or reverse remodeling of the myocardium need further elucidation. To accomplish this, accurate and reliable characterization of the myocardium is crucial. Multiparametric cardiovascular magnetic resonance (CMR) imaging uniquely enables measurement of cardiac structure and function as well as tissue characterization. Late gadolinium enhancement (LGE) offers direct visualization of replacement myocardial fibrosis while T1 mapping quantitatively assesses diffuse interstitial myocardial fibrosis, both of which are hallmark of adverse myocardial remodeling. Performed at different disease states and at planned time points, CMR could provide insights into cardiac remodeling disease biology and prognosis among individuals with diverse cardiometabolic profile. In addition, the inherent power of magnetic resonance imaging (MRI) in discerning fat and water signals makes it ideal for fat visualization and quantification across anatomical locations key in cardiometabolic disorders. Meanwhile, the examination of surrogate biomarkers complementary to imaging features supplements mechanistic understanding and offers translational opportunities for health or disease monitoring.

The REMODEL (Response of the Myocardium to Hypertrophic Conditions in the Adult Population) study aims to characterize and establish the significance and mediators of myocardium remodeling underlaid by cardiometabolic risks, enabled by multiorgan, multiparametric MRI and biochemical markers.

## METHODS

### Study Design and Population

REMODEL is a prospective observational cohort study that started in 2016 to study the role of CMR in patients with hypertension. In 2024, REMODEL has expanded beyond hypertension to include patients with one or more of the following cardiometabolic risk factors namely hypertension, diabetes mellitus, dyslipidemia, fatty liver disease and obesity. Participants are recruited from outpatient clinics at National Heart Centre Singapore and primary care setting, as well as members of the public through approved advertisements.

All participants provide written informed consent to follow-up and data collection for 10 years. A subset of participants who consent to follow-up CMR imaging are re-scanned to study disease progression. All participants will have clinical outcome adjudicated. The most current eligibility criteria are detailed in **Table 1**.

**Table 1.**
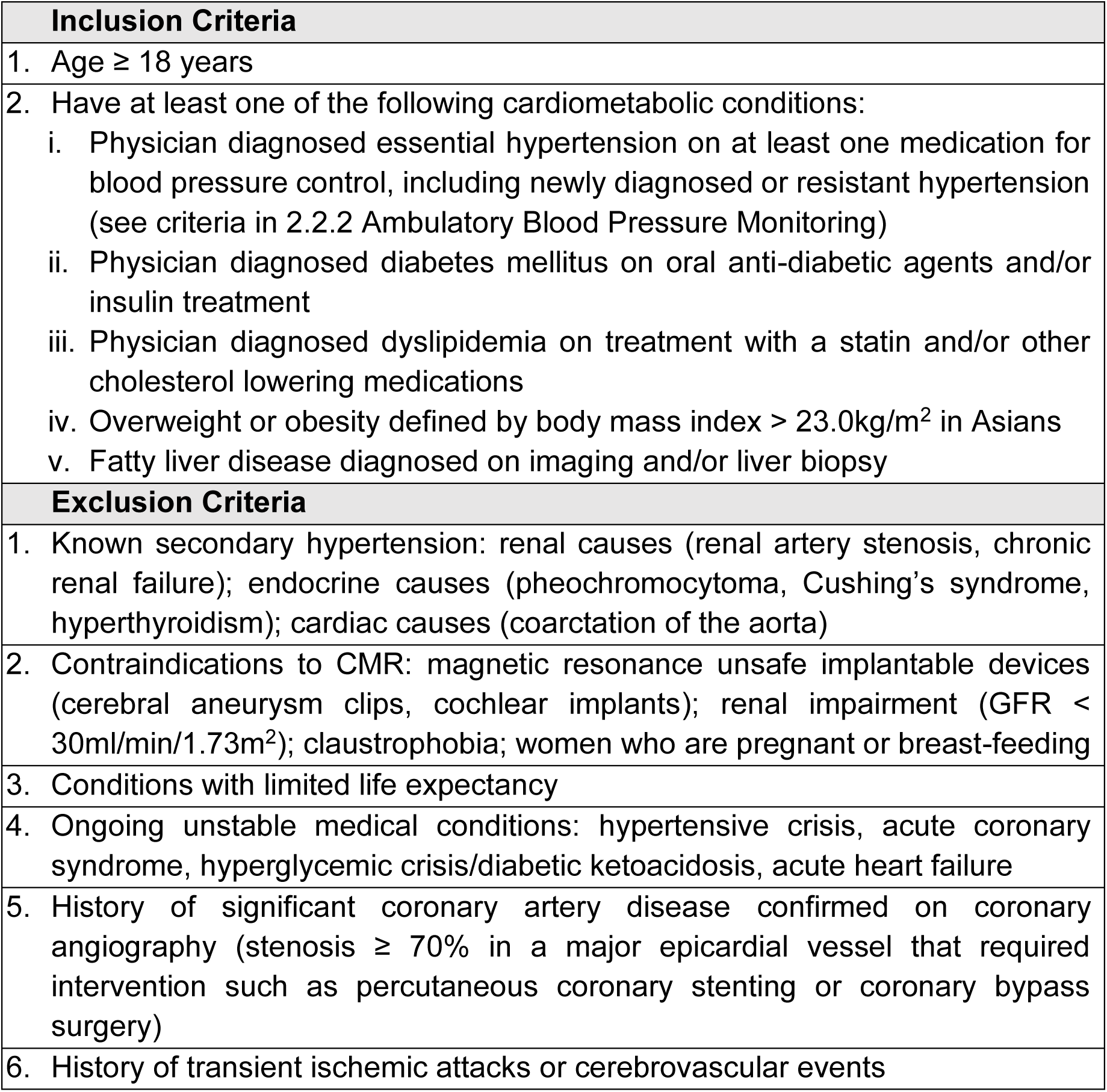
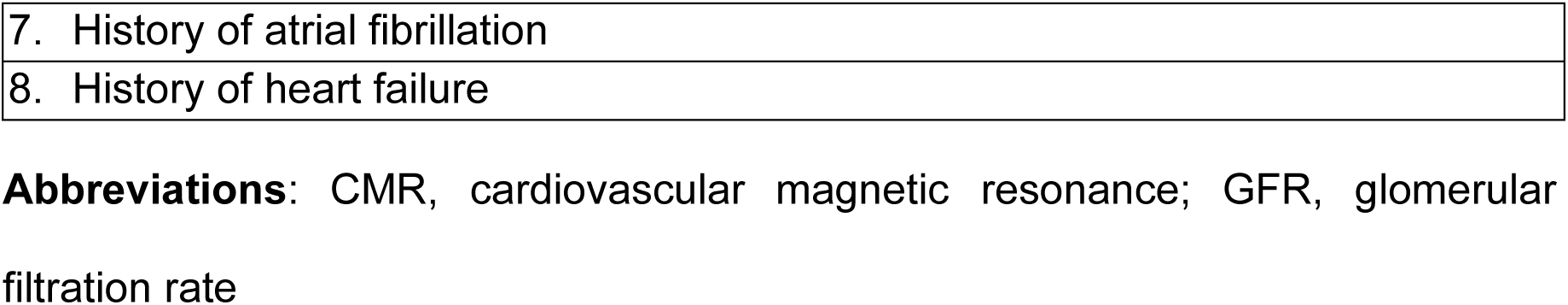
Eligibility criteria of REMODEL.

| **Inclusion Criteria** |
| --- |
| 1. Age ≥ 18 years |
| 2. Have at least one of the following cardiometabolic conditions:<br>i. Physician diagnosed essential hypertension on at least one medication for blood pressure control, including newly diagnosed or resistant hypertension (see criteria in 2.2.2 Ambulatory Blood Pressure Monitoring)<br>ii. Physician diagnosed diabetes mellitus on oral anti-diabetic agents and/or insulin treatment<br>iii. Physician diagnosed dyslipidemia on treatment with a statin and/or other cholesterol lowering medications<br>iv. Overweight or obesity defined by body mass index $> 23.0\text{kg/m}^2$ in Asians<br>v. Fatty liver disease diagnosed on imaging and/or liver biopsy |
| **Exclusion Criteria** |
| 1. Known secondary hypertension: renal causes (renal artery stenosis, chronic renal failure); endocrine causes (pheochromocytoma, Cushing's syndrome, hyperthyroidism); cardiac causes (coarctation of the aorta) |
| 2. Contraindications to CMR: magnetic resonance unsafe implantable devices (cerebral aneurysm clips, cochlear implants); renal impairment ($\text{GFR} < 30\text{ml/min/1.73m}^2$); claustrophobia; women who are pregnant or breast-feeding |
| 3. Conditions with limited life expectancy |
| 4. Ongoing unstable medical conditions: hypertensive crisis, acute coronary syndrome, hyperglycemic crisis/diabetic ketoacidosis, acute heart failure |
| 5. History of significant coronary artery disease confirmed on coronary angiography (stenosis $\geq 70\%$ in a major epicardial vessel that required intervention such as percutaneous coronary stenting or coronary bypass surgery) |
| 6. History of transient ischemic attacks or cerebrovascular events |
| 7. History of atrial fibrillation |
| 8. History of heart failure |

**Abbreviations:** CMR, cardiovascular magnetic resonance; GFR, glomerular filtration rate

### Study Procedures and Investigations

Overview of study procedures from recruitment to baseline and follow-up investigations are depicted in **Figure 1**. Minimally, consent to baseline data collection, CMR and blood sampling is required for inclusion.

**Figure 1.**
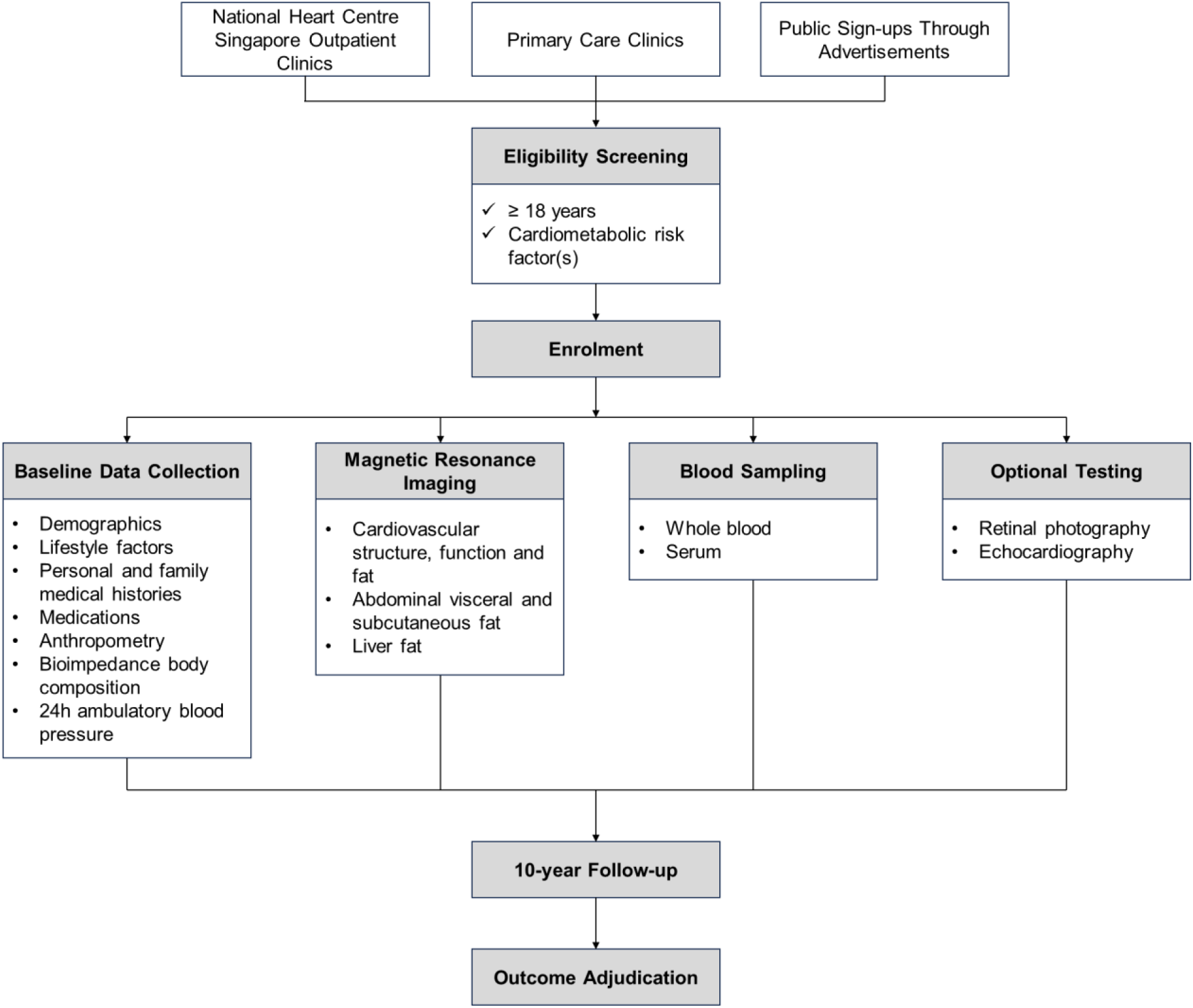
REMODEL study procedure. *Performed in consenting participants.

#### Demographic, Clinical and Anthropometric Data

At enrolment, demographic particulars (age, sex, ethnicity), lifestyle factors (alcohol consumption, smoking habit), past medical history, family medical history, presence and duration of current medical conditions, and current medications are documented. Height, weight, waist circumference and hip circumference are measured using standard method in a standing position. Waist circumference is measured at just above the navel. Hip circumference is taken at the widest portion of the hip area.

Body composition is measured using the InBody 370 bioimpedance analyzer (InBody, Cerritos, California, USA) that reports body fat mass, skeletal muscle mass and fat-free masses of water, protein and minerals.

#### Ambulatory Blood Pressure Monitoring

mbulatory blood pressure monitoring is administered for participants with existing hypertension. A correctly sized cuff of the OnTrak 90277 device (Spacelabs Healthcare, Snoqualmie, WA, USA) is applied to the participant’s non-dominant arm. Over 24 hours, automated out-of-clinic blood pressure measurements are taken every 20 minutes between 6am-10pm, and every 30 minutes between 10pm-6am the following day.

Hypertension on ambulatory blood pressure monitoring is defined as systolic blood pressure ≥ 135 mmHg (day), ≥ 120 mmHg (night) or ≥ 130 mmHg (24-hour); and diastolic blood pressure ≥ 85 mmHg (day), ≥ 70 mmHg (night) or ≥ 80 mmHg (24-hour)[6].

#### Cardiovascular Magnetic Resonance (CMR)

CMR is performed in all participants for evaluation of cardiac volumes, structure and function.

##### Image Acquisition

CMR scans are performed on a MAGNETOM Aera 1.5T scanner (Siemens Healthineers, Erlangen, Germany). A standardized scanning protocol is used: long-axis balanced steady-state free precession cine images are acquired in the two-, three-, and four-chamber views; and short-axis cines are acquired from the mitral valve annulus to the apex (acquired voxel size 1.6 × 1.3 × 8.0 mm; 30 phases per cardiac cycle).

Non-ischemic focal replacement fibrosis and diffuse interstitial myocardial fibrosis are evaluated with LGE and T1 mapping techniques, respectively. LGE is performed 8 minutes after contrast administration of 0.1 mmol/kg of gadobutrol (Gadovist; Bayer Pharma AG, Germany). A phase-sensitive inversion-recovery (PSIR) fast gradient echo sequence is used, and the inversion time is optimized to achieve appropriate nulling of the myocardium. T1 mapping is performed using the modified Look-Locker inversion-recovery sequence. Native T1 map is acquired using a heartbeat scheme of 5(3)3; post-contrast T1 map is acquired 15 minutes after contrast administration using a heartbeat scheme of 4(1)3(1)2.

In addition to CMR, liver fat and abdominal fat imaging have been added in 2018 and 2021, respectively. Imaging protocol has been published[5]. Measurement of liver fat, expressed as proton density fat fraction (PDFF), is performed using the Liver*MultiScan*-Iterative Decomposition of water and fat with the Echo Asymmetry and Least Squares estimation method (*LMS IDEAL*; Perspectum Ltd, Oxford, London, UK). This approach has been verified across MRI platforms and has accuracy within 3% of lab-analyzed fat samples[7]. The sequence parameters are TEs: 1.30, 3.30, 5.30, 7.30, 9.30 and 11.30 ms; TR: 14 ms; flip angle: 5 degrees; matrix size: 232 × 256mm^2^; FOV: 398 × 440 mm^2^; slice thickness: 10mm; number of slices: 5; slice gaps: 5 mm.

Abdominal visceral adipose tissue (VAT) and subcutaneous adipose tissue (SAT) are quantified from contiguous short-axis fat– and water-separation images from the two-point Dixon method. The sequence parameters are TEs: 2.39 and 4,77 ms; TR:

6.5 ms; flip angle: 10 degrees; matrix size: 260 × 320 mm^2^; FOV: 325-366 × 400-450 mm^2^; slice thickness: 4 mm; slice gap: 0.8 mm[8].

##### Image Analysis

All images are de-identified and analyzed at the National Heart Research Institute (NHRIS) CMR Core Lab by trained personnel blinded to participants’ data.

CMR analyses are performed using standardized protocols using cvi42 image analysis software (Circle Cardiovascular Imaging, Calgary, Canada)[9,10]. Left ventricular (LV) mass (g) and cardiac volumes (mL) are indexed to body surface area (m^2^) by the Du Bois method. Population-specific sex– and age-corrected reference ranges are used for LVH classification[9]. Ejection fractions (%) are calculated as percentage of stroke volume over end-diastolic volume.

LV concentricity is defined as the ratio between LV mass and end-diastolic volume (EDV)[11]. The Remodeling Index, which is inversely correlated with global myocardial wall stress, is derived as 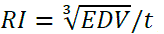, where EDV is the LV end-diastolic volume (mL) and *t* is the maximal wall thickness (cm) across 16 myocardial segments[10,12]. Multidirectional LV strain analysis includes global radial strain (GRS), global longitudinal strain (GLS) and global circumferential strain (GCS). Better strain function corresponds with larger GRS values and conversely with smaller (more negative) GLS and GCS values.

Myocardial fibrosis is examined as two phenotypes: diffuse interstitial fibrosis denoted by interstitial volume, and replacement fibrosis qualified by LGE (**Figures 2A, B**). Interstitial volume (mL) is the product of extracellular volume (ECV) fraction × myocardial volume (mL), where myocardial volume = absolute LV mass/specific gravity of myocardium, which is 1.05 g/mL. Presence of LGE is stratified into ischemic and non-ischemic fibrosis.

**Figure 2.**
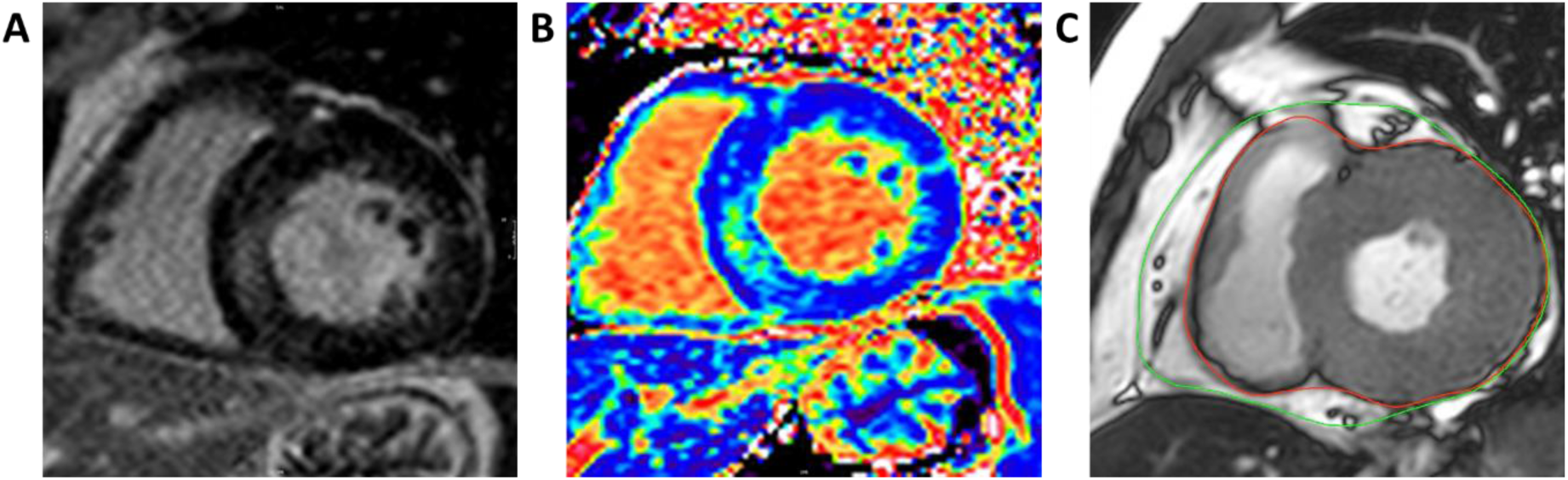
Cardiovascular magnetic resonance imaging for assessment of myocardial fibrosis and epicardial fat. **(A)** Late gadolinium enhancement and **(B)** extracellular volume mapping in a patient with hypertension and diabetes mellitus. **(C)** Epicardial fat contour between the epicardium (green) and pericardium (red).

Epicardial adipose tissue (EAT) is quantified as volume (cm^3^) using the modified Simpson’s rule by summation of short-axis stack from the mitral valve annulus to the apex covering both left and right ventricles. In each end-systolic short axis image, EAT is delineated as the fat layer between the myo-epicardium and the pericardium, excluding the paracardial fat which surrounds the external margin of the pericardium[5] (**Figure 2C**).

Liver PDFF (%) is computed as fat/(fat+water) in the regions of interest on the PDFF parametric map, avoiding image artifacts and vessels[13,14]. Transversal areas of VAT and SAT (cm^2^) are segmented from a single-slice image at the L3 vertebral level using ITK-SNAP software version 3.8 (PICLS, University of Pennsylvania, USA). This approach has been verified against the total VAT and SAT volume in men and women (correlation, r > 0.9)[15,16].

#### Biochemical Markers

Serum and whole blood samples are collected and stored at –80°C at the National Heart Centre Singapore Biobank until analysis. Follow-up serum samples are collected in the subset of participants who consent to re-imaging.

#### Other Procedures

Consenting participants are invited to other related imaging studies including retinal photography and echocardiography.

#### Outcome Measures

Adverse events accounted for include hospitalizations for heart failure, unstable angina, stroke, myocardial infarction and mortality. The cause of death is obtained from the National Death Registry.

### Data Analysis Plan

The association of clinical and metabolic determinants (from preexisting conditions or newly mapped liver and abdominal adiposity) with CMR features of cardiac remodeling will be determined. Levels of circulating biomarkers and proteomic expressions by metabolic status will be examined in the context of morphological and functional findings on CMR for mechanistic elucidation. Disease progression and prognosis by cardiometabolic health and remodeling condition will be assessed using follow-up and adjudicated outcome data.

Continuous variables that are normally and non-normally distributed will be presented as mean ± standard deviation (SD) and median (interquartile range), respectively. Frequency of categorical data will be described as number (percentage). Missing data will be censored and not be imputed. Group comparisons will be conducted using the Student’s t-test, Mann-Whiney U test or χ^2^ test accordingly. Multivariable linear or logistic regressions adjusted for clinically important covariates will be performed to determine independent associations between variables. Subgroup analyses will be examined with the analysis of covariance (ANCOVA). Event-free survival analysis will be estimated using the Kaplan-Meier curve, compared with the log-rank test and modelled with covariates using the Cox proportional hazards model. Statistical significance is defined as 2-sided P<0.05. Statistical analyses will be performed using IBM SPSS Statistics Version 26 (IBM Corp, Armonk, NY, USA) and GraphPad Prism Version 7.05 (GraphPad Software, Inc, La Jolla, CA, USA).

### Data Management

Physical study binders are secured in locked cabinets in a locked office at National Heart Centre Singapore. Access is limited to the principal investigator, data manager, clinical research coordinators and selected research team members. Clinical data, MRI images, biological specimens are anonymized to ensure participants’ confidentiality and analysis blinding. Data are curated in an in-house database.

### Ethics and Dissemination

The study is performed in accordance with ethical principles that have their origin in the Declaration of Helsinki. Ethics and protocol amendment approvals were granted by the SingHealth Centralized Institutional Review Board (2015/2603). The study is registered in ClinicalTrials.gov (NCT02670031). All participants receive detailed explanation of the study details pertaining to risks and benefits by trained clinical research coordinators before providing written informed consent. Participation and withdrawal are voluntary and do not affect standard clinical care. Data will be anonymized and aggregated for analysis, reporting and publication.

## RESULTS

### Study Population

At the time of reporting, more than 1,000 participants have been recruited. All participants have baseline CMR and blood samples while more than 400 have retinal photography done. Repeat CMR is available in 200 follow-ups. Participants’ baseline characteristics are shown in **Table 2**.

**Table 2.**
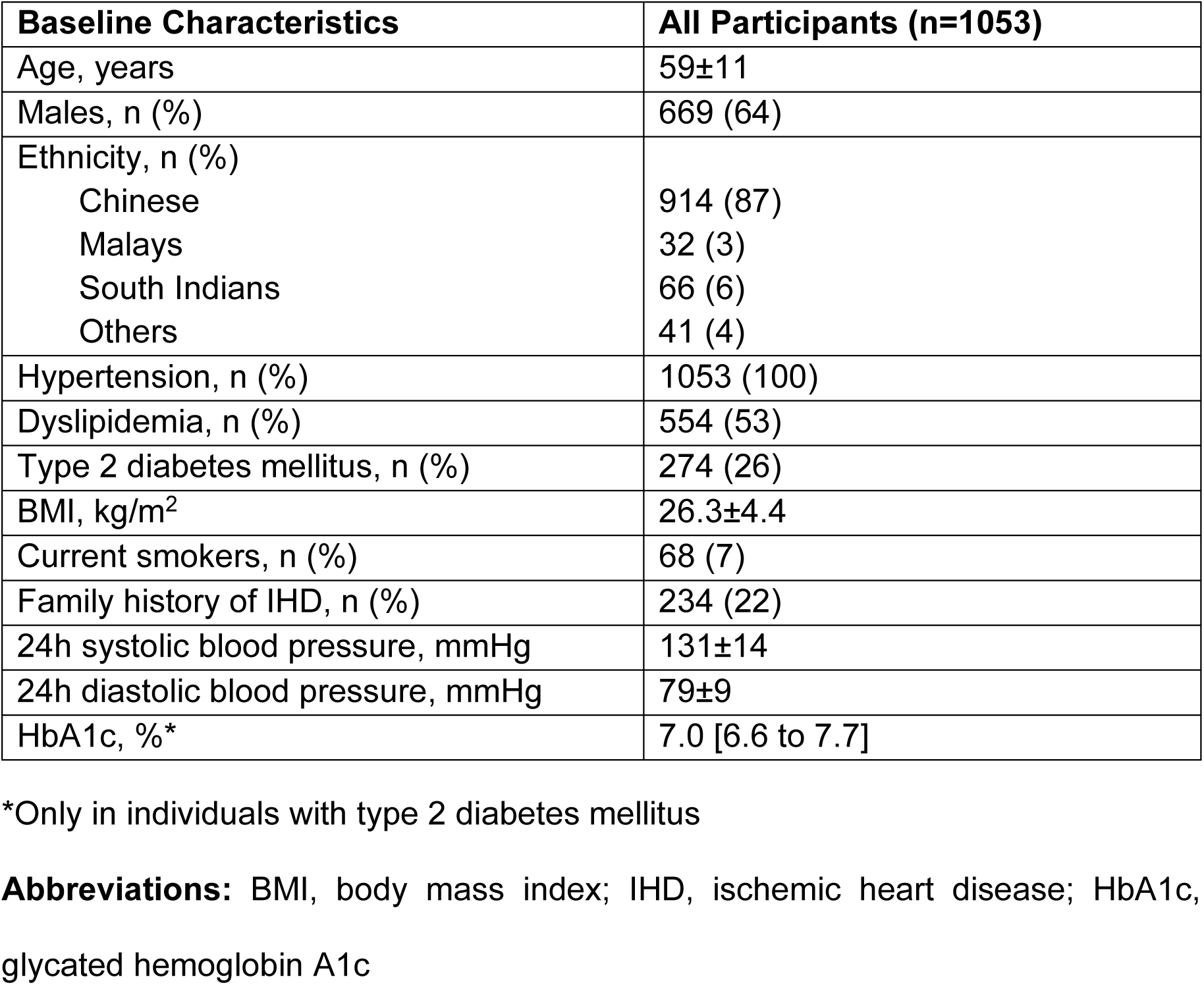
Baseline characteristics of study participants to-date.

### Study Outputs and Collaborations

REMODEL has contributed to multi-disciplinary collaborations and knowledge advancement pertaining to method development, disease biology, diagnosis and prognosis (**Online Supplemental Table 1**). In 2019, a randomized controlled trial was incorporated in the REMODEL program, randomizing patients with hypertensive LVH to either sacubitril/valsartan or valsartan (NCT03553810)[17]. The primary objective was to compare the efficacy of the two medications in reducing myocardial fibrosis on CMR. In 2022, REMODEL further strengthened its efforts in precision medicine by partnering with RESET, a program funded by the National Medical Research Council, to explore the immune, lipid and metabolic mediators of early cardiovascular disease (NCT06211868). Additionally, on-going multi-omics analyses of participants in REMODEL are contributing to national initiatives in precision medicine, enabling more personalized approaches to cardiovascular care.

## DISCUSSION

REMODEL is positioned to be a prominent prospective observational cohort study using CMR to understand the significance of cardiac remodeling in individuals with cardiometabolic disease. The study draws strengths from several aspects: substantial sample size, demographic relevance to South-East Asian population, prospective and longitudinal investigation, comprehensive CMR characterization, and biomarker integration.

Multiparametric CMR allows accurate characterization of the myocardium and delineation of disease phenotypes. Efficiency and reproducibility of contouring and measurement of CMR metrics have improved with scan resolution enhancement and incorporation of machine learning in post-scan analysis. A key and unique strength of CMR lies in the myocardial T1 mapping feature which permits direct visualization and quantification of diffuse myocardial fibrosis, which is an early, reversible hallmark of myocardial remodeling in heart failure. Being a non-invasive modality, repeat CMR assessments enable the study of disease progression at no ionizing radiation risk to participants.

Distinct from previous studies that are confined in phenotypes of certain metabolic disorder or cardiac dysfunction, REMODEL recognizes the inseparable nature of metabolic risk factors such that its holistic approach will offer valuable insights with clinical relevance. Furthermore, there will be opportunities to compare the disease cohort with matched healthy cohort for further understanding of the pathological implication of cardiometabolic disease, paving the way for future studies and disease monitoring strategies. Indeed, REMODEL’s demographic uniqueness, data comprehensiveness and quality have proved to be invaluable in past and ongoing multi-disciplinary collaborations. In the translational aspect, CMR-verified biochemical markers will be of practical and economical value in healthcare implementation.

## Data Availability

All data produced in the present work are contained in the manuscript

## ACKNOWLEDGEMENT

The authors wish to thank all study participants, referring physicians, and radiographers at the Department of Cardiovascular Magnetic Resonance, National Heart Centre Singapore for their support in the study.

## AUTHORS’ CONTRIBUTIONS

VL, TTL and CWLC contributed to the conception and design of the study. JAB, YH and RB contributed to the acquisition and analysis of the data for the work. ABL, WT, SCK and DFT contributed to patient recruitment, collection and curation of data. VL and CWLC drafted the manuscript. TP, MSLK, CHL, TTL and CWLC made critical revisions to the manuscript. All authors gave final approval and agree to be accountable for all aspects of work ensuring integrity and accuracy.

## PATIENT AND PUBLIC INVOLVEMENT

Research findings are regularly shared with patients and the public through social media, traditional news outlets and public forums.

## SUPPLEMENTAL MATERIAL

Online Supplemental Table S1

