## Supplementary material for "Response of the Myocardium to Hypertrophic Conditions in the Adult Population: The Prospective Observational REMODEL Cohort Study Protocol": Online Supplemental Table S1

**Online Supplemental Table 1.** Publications to-date resulting from the REMODEL population.

| **Author** | **Main Findings** |
|---|---|
| Chin 2017[18] | Editorial on targeting the myocardium in hypertensive LVH. |
| Goh *et al.* 2017[10] | The novel Remodeling Index is derived as a marker of advanced hypertrophy in patients with hypertensive LVH. |
| Lim *et al.* 2018[19] | Sex-specific regression model improves estimation of synthetic hematocrit and extracellular volume in hypertensive patients. |
| Chua *et al.* 2019[20] | Retinal capillary density is associated with higher blood pressure and lower eGFR. |
| Chua *et al.* 2019[21] | Elevated blood pressure is associated with increased choriocapillaris flow voids on optical coherence tomography angiography in hypertensive patients. |
| Chua *et al.* 2021[22] | Choriocapillaris microvasculature dysfunction is associated with poor blood pressure control and renal dysfunction. |
| Le *et al.* 2021[12] | The Remodeling Index provides prognostic value that improves risk stratification of hypertensive LVH. |
| Le *et al.* 2021[23] | Echocardiographic left ventricular global longitudinal strain is associated adverse cardiac remodeling on CMR and confers worse prognosis. |
| Chua *et al.* 2022[24] | Altered retinal capillary density is correlated with adverse cardiac remodeling on CMR. |
| Iyer *et al.* 2022[25] | Hypertensive myocardial fibrosis is associated with adverse cardiac remodeling and outcomes. |
| Pua *et al.* 2023[4] | Hypertension coexisting with diabetes confers worse adverse cardiac remodeling, associating with markedly increased proteomic expressions of immune and inflammatory response. |
| Lee *et al.* 2023[17] | Protocol of the prospective, randomized, open-label, blinded endpoint trial, REVERSE-LVH. |
| Lee *et al.* 2024[5] | Abdominal visceral, liver and epicardial fats are associated with adverse cardiac remodeling. |

**Abbreviations:** CMR, cardiovascular magnetic resonance; eGFR, estimated glomerular filtration rate; LVH, left ventricular hypertrophy.
